# Association between elevated Lp(a) and CAV in heart transplant recipients

**DOI:** 10.1101/2025.05.26.25328315

**Authors:** Lin Cao, Shi Huang, Kaushik Amancherla, Sandip K. Zalawadiya, Hasan K. Siddiqi, Kelly Schlendorf, JoAnn Lindenfeld, Wen-Liang Song

## Abstract

**Background:** Cardiac allograft vasculopathy (CAV) is the leading cause of late graft failure and mortality after heart transplantation (HT). Among non-HT patients, elevated Lp(a) levels are associated with increased atherosclerotic cardiovascular disease (ASCVD) risk. However, it is less clear what role Lp(a) may play in the development of CAV and how it interacts with other lipids post-transplant.

**Objectives:** We sought to examine the association between elevated Lp(a) and CAV among adult HT recipients at a single high-volume heart transplant center.

**Methods:** We performed a retrospective analysis of 504 adult HT recipients followed at the transplant center who had undergone coronary angiography for CAV evaluation within 5 years post-transplant. Maximum Lp(a) values and lipid panels obtained concurrently were collected. Patients were considered to have CAV if they had CAV 1 or higher by ISHLT grading on any coronary angiography obtained within 5 years post-transplant.

**Results:** In our cohort, 114 patients developed CAV. Among all patients who developed CAV, there was a significant difference in 5-year incidence rate of CAV when stratified by Lp(a) ≥ 90 mg/dL or < 90 mg/dL. The incidence of CAV was 9.4 per 100 person/years in the lower Lp(a) group versus 15.2 per 100 person/years in the higher Lp(a) group (p=0.031). Kaplan-Meier plots of CAV-free survival showed a significant association between CAV-free probability and Lp(a) ≥ 90 mg/dL (p=0.037). After stratifying Kaplan-Meier plots by LDL, the relationship between Lp(a) ≥ 90 mg/dL and CAV-free probability remained significant (p=0.012) among patients with LDL ≤ 100 mg/dL but was no longer significant (p=0.85 in patients with LDL > 100 mg/dL. Among patients with LDL ≤ 100 mg/dL, the Cox model results suggested higher Lp(a) levels increase the risk of developing CAV after HT (HR 1.35, 95% CI 1.03-1.76, p=0.03).

**Conclusions:** Among post-HT patients, Lp(a) ≥ 90 mg/dL was associated with increased risk for development of CAV, particularly at lower LDL levels. In patients with LDL ≤ 100 mg/dL, Lp(a) ≥ 90 mg/dL was associated with CAV-free survival. Lp(a) may be a useful biomarker for risk stratification in transplant patients and a potential future target for CAV prevention with emerging Lp(a) therapeutics.

## Introduction

Cardiac allograft vasculopathy (CAV) remains a leading cause of morbidity and mortality after heart transplantation (HT) despite newer approaches for prevention and management^1,2^. Approximately 10% of HT recipients develop CAV within 1-year post-transplant, and over half of all recipients are diagnosed with CAV by 10-years post-transplant^1^. CAV is unique to transplanted hearts and can present in a variety of ways with lesions ranging from diffuse concentric intimal fibromuscular hyperplasia to plaques more classically resembling atherosclerosis in the epicardial and branch vessels and microvasculature^3,4^. Vascular injury is thought to be mediated by chronic endothelial damage and repair leading to hyperproliferation of smooth muscle and connective tissue with eventual lumen compromise^3,4^. Immune and inflammatory factors such as ischemia-reperfusion injury, allograft immune response, and viral infections including cytomegalovirus (CMV) all contribute, along with traditional cardiac disease risk factors^3^.

Dyslipidemia is common post-transplant and is a known risk factor for CAV^3,5^. Prior work has found that elevated cholesterol, particularly low-density lipoprotein (LDL), damages the endothelium, induces fibrotic proliferative changes, and impairs essential functions like vasorelaxation^3,6^. In addition, examination of arteries affected by CAV often reveals both intra- and extracellular lipid accumulation within the subendothelial tissue, and the degree of lipid accumulation is correlated with luminal narrowing^6^. Early initiation of statins post-transplant has been associated with improved post-transplant dyslipidemia, increased one-year survival, decreased rejection, and decreased incidence of CAV^5,7^. Statin therapy is a class 1 level A recommendation in the 2023 International Society of Heart and Lung Transplantation guidelines for all HT adult and pediatric recipients regardless of lipid levels^7^.

More recently, lipoprotein(a) [Lp(a)] has emerged as a target for prevention of adverse cardiac events in the general population. Lp(a) is an LDL-like particle whose levels are predominantly genetically determined, and it consists of an apoB100-containing lipoprotein bound to apolipoprotein(a)^8^. In the general population, Lp(a) has been strongly, causally associated with higher atherosclerotic cardiovascular disease risk, with increasing risk with higher Lp(a) levels^9,10,11,12^. This relationship holds true across all levels of LDL-C and apoB^9,13^. A variety of pathophysiological mechanisms have been proposed. In addition to having the atherogenic LDL-like apo-B component, the apo(a) subpart stimulates proliferation of smooth muscle cells and increases binding with arterial walls, likely through its interactions with plasmin and transforming growth factor-β^14,15,16^. Lp(a) also associates with inflammatory oxidized phospholipids and can serve as an immune cell activator^14,16,17^.

However, it remains less clear if Lp(a) contributes to the development of CAV given that the underlying pathophysiology differs from atherosclerosis in the general population in several ways^6^. Uncertainties also remain about how Lp(a) mediated CAV risk might interact with CAV risk due to other lipids post-transplant. Several small studies thus far have suggested that elevated Lp(a) levels are fairly prevalent in HT patients and may contribute to development of CAV^18,19^. We sought to examine the association between Lp(a) and CAV among HT recipients at a single large-volume HT center.

## Methods

We conducted a retrospective chart review of adult HT recipients transplanted at our center who had Lp(a) levels collected pre- or post-transplant and had undergone coronary angiography for CAV evaluation. If patients had undergone multiple transplants, only their first transplant was included. If patients had multiple Lp(a) values, the maximum Lp(a) value was used. Patients were considered to have CAV if they had CAV 1 or higher by ISHLT grading on any coronary angiography obtained within 5-years post-transplant. Lipid panels obtained at the same time as the Lp(a) values were also collected. If no simultaneous lipid panel was available, the lipid panel drawn most close in time to the Lp(a) value was used. Donor age, recipient tobacco use history, and presence of hypertension and diabetes at HT based on diagnostic codes and provider notes were also collected.

### Statistical Analysis

Descriptive statistics (i.e., median (IQR), n (%)) for demographic and clinical characteristics are presented. Wilcoxon and Chi-squared tests were used to examine differences in age at transplant, transplant indication, sex, donor age, recipient tobacco use history, presence of hypertension, presence of diabetes, Lp(a), and LDL levels between patients who developed CAV and patients who did not. A cutoff of Lp(a) 90 mg/dL was utilized. While there are no distinct cutoffs for defining elevated Lp(a) levels and increasing levels are associated with increasing ASCVD risk, Lp(a) >90 mg/dL represents the top 5% of Lp(a) concentrations in the Copenhagen General Population and Copenhagen City Heart studies and some have proposed 90 mg/dL as a threshold for extremely high risk^20^. In addition, trials examining Lp(a) therapeutics have used Lp(a) cutoffs of 90 mg/dL (HORIZON NCT04023552). The 5-year incidence rate of CAV was calculated, and rate ratios and 95% confidence intervals between patients with Lp(a) < 90 mg/dL and Lp(a) ≥ 90 mg/dL.

A Kaplan-Meier plot was used to present probabilities of freedom from CAV events stratified by patients with Lp(a) ≥ 90 mg/dL vs Lp(a) < 90 mg/dL and separated Kaplan-Meier plots were created stratified by patients’ LDL levels (i.e., LDL ≤ 100 mg/dL and LDL > 100 mg/dL). Cox proportional hazards models were constructed to examine how Lp(a) levels impact the risk of developing CAV after HT. The Cox models adjusted for potential confounders such as age at transplant, donor age, sex, tobacco use history, hypertension status, diabetes status, transplant indication, and LDL levels. In addition, separate Cox models were constructed by LDL levels using a cutoff of 100 mg/dL. All statistical analyses were performed using R (version 4.4.0).

## Results

Among 504 adult HT patients included in our study (median follow-up 1.12 years), 390 patients remained CAV free (median follow-up 1.46 years) and 114 developed CAV (median follow-up 1.01 years). Patients who remained CAV free tended to have been followed longer (p<0.001). There were no significant differences between the two groups in age at transplant, sex, transplant indication, rates of hypertension, or rates of diabetes. Difference in recipient tobacco use history was significant (p=0.021), with a greater proportion of patients who developed CAV having a smoking history (57.9% vs 45.6%). Patients who developed CAV tended to have been transplanted using older donors (33.49 ± 10.02 vs 29.13 ± 9.59 years, p=<0.001). Average maximum Lp(a) levels were similar between the two groups (53.73 ± 51.71 vs 45.53 ± 47.24 mg/dL in CAV positive and CAV free groups respectively, p=0.383). There was a significant difference in proportion of patients with Lp(a) ≥ 90 mg/dL (26.3% vs 17.4% in the CAV positive and CAV free groups respectively, p=0.035). There was no significant difference in LDL levels between the two groups (73.2 ± 32.3 vs 76.1 ± 29.7 mg/dL in the CAV positive and CAV free groups respectively, p=0.265). (Table 1).

**Table 1.** Descriptive statistics, ischemic cardiomyopathy (ICM), nonischemic cardiomyopathy (NICM)

|  | **CAV free**<br>**(390 patients)** | **Developed CAV**<br>**(114 patients)** | **P-**<br>**value** |
| --- | --- | --- | --- |
| **Age at transplant** | 52.9 [43.0-62.5]<br>51.8 ± 13.1 | 54.1 [45.2-62.6]<br>52.5 ± 12.5 | 0.707 |
| **Donor age** | 28.00 [21.00-35.00]<br>29.13 ± 9.59 | 33.00 [27.00-41.00]<br>33.49 ± 10.02 | <0.001 |
| **Sex** |  |  | 0.06 |
| **Female** | 31.0% (121) | 21.9% (25) |  |
| **Male** | 69.0% (269) | 78.1% (89) |  |
| **Transplant indication** |  |  | 0.828 |
| **ICM** | 32.1% (125) | 35.1% (40) |  |
| **NICM** | 64.1% (250) | 61.4% (70) |  |
| **Mixed** | 3.8% (15) | 3.5% (4) |  |
| **Recipient tobacco use history** |  |  | 0.021 |
| **Yes** | 45.6% (178) | 57.9% (66) |  |
| **No** | 54.4% (212) | 42.1% (48) |  |
| **Hypertension** |  |  | 0.134 |
| **Yes** | 84.1% (328) | 78.1% (89) |  |
| **No** | 15.9% (62) | 21.9% (25) |  |
| **Diabetes** |  |  | 0.09 |
| **Yes** | 50.3% (196) | 41.2% (47) |  |
| **No** | 49.7% (194) | 58.8% (67) |  |
| **Time from transplant to CAV development or total follow-up (years)** | 1.458 [0.988-4.945]<br>3.648 ± 4.705 | 1.005 [0.655-1.797]<br>1.345 ± 1.110 | <0.001 |
| **LDL (mg/dL)** | 73.0 [57.0-91.0]<br>76.1 ± 29.7 | 67.0 [49.5-91.0]<br>73.2 ± 32.3 | 0.265 |
| **Max Lp(a) (mg/dL)** | 25.00 [11.00-69.00]<br>45.53 ± 47.24 | 33.50 [9.25-93.75]<br>53.73 ± 51.71 | 0.383 |
| **Categorical Lp(a)** |  |  | 0.035 |
| **< 90 mg/dL** | 82.6% (322) | 73.7% (84) |  |
| **≥ 90 mg/dL** | 17.4% (68) | 26.3% (30) |  |

The incidence of CAV was 10.5 per 100 person/years among all patients. There was a significant difference in 5-year incidence rate of CAV when stratified by Lp(a) ≥ 90 mg/dL or < 90 mg/dL. The incidence of CAV was 9.4 per 100 person/years in the Lp(a) <90 mg/dL group versus 15.2 per 100 person/years in the Lp(a) ≥90 mg/dL (p=0.031).

A Kaplan-Meier plot of CAV-free probability stratified by Lp(a) ≥ 90 mg/dL and Lp(a) < 90 mg/dL is shown in **figure 1**. There was a significant difference in CAV-free probability between the two groups (p=0.037). The relationship between Lp(a) and CAV was examined at various LDL levels as well. Separated Kaplan-Meier plots in patients with LDL ≤ 100 mg/dL and LDL > 100 mg/dL are shown in **figure 2a and b**. The difference in CAV-free probability was significant between the two groups in patients with LDL ≤ 100 mg/dL (p=0.012) and was no longer significant in patients with LDL > 100 mg/dL (p=0.85).

**Figure 1.**
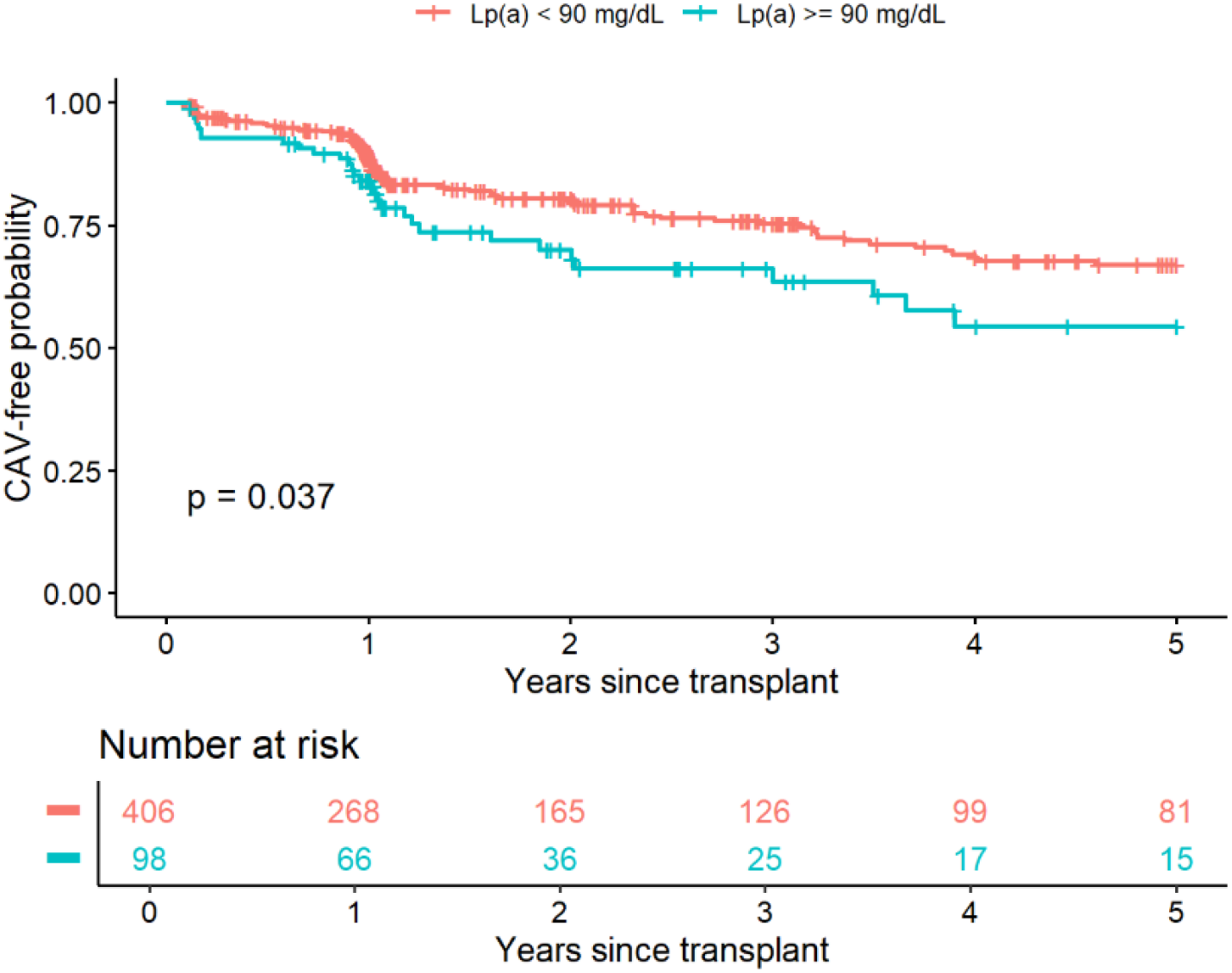
Kaplan-Meier plot of CAV free probability stratified by Lp(a) ≥ 90 mg/dL or < 90 mg/dL

**Figure 2a and b.**
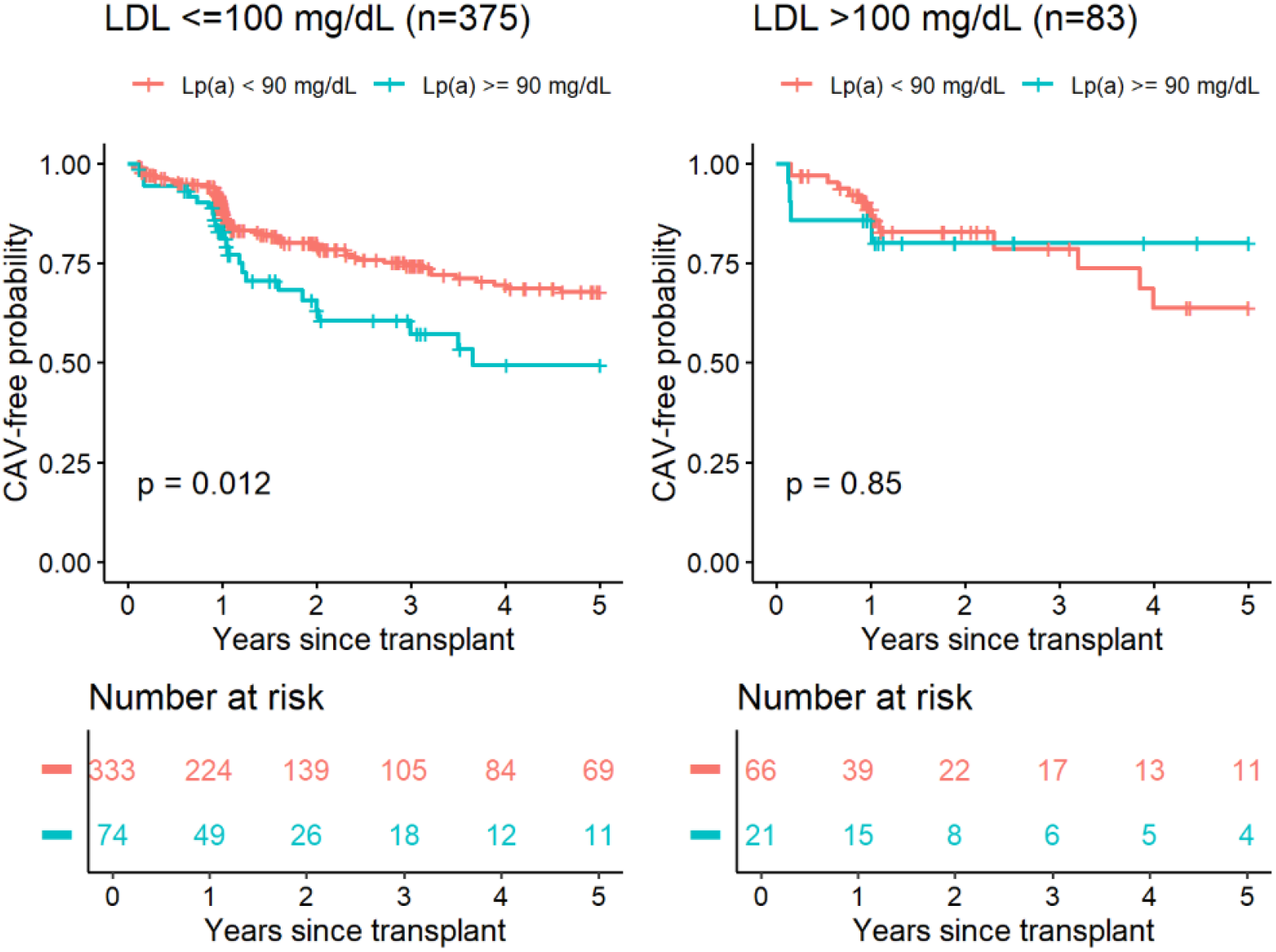
Kaplan-Meier plot of CAV free probability by LDL stratified by Lp(a) ≥ 90 mg/dL or < 90 mg/dL

A Cox regression adjusting for potential confounders (age at transplant, sex, LDL levels, history of tobacco use, presence of hypertension, presence of diabetes, donor age, and transplant indication) demonstrated no significant relationship between Lp(a) and CAV with Lp(a) 29 mg/dL vs 90 mg/dL (HR 1.18, 95% CI 0.94-1.49). Among patients with LDL ≤ 100 mg/dL, the Cox model results suggested higher Lp(a) levels increase the risk of developing CAV after HT (HR 1.35, 95% CI 1.03-1.76, p=0.03). The model-estimated cumulative CAV incidence at five years by Lp(a) levels is presented in **Figure 3**. However, the relationship between Lp(a) levels and risk of developing CAV was not significant among patients with LDL >100 mg/dL (HR 0.68, 95% CI 0.34-1.34, p=0.26).

**Figure 3.**
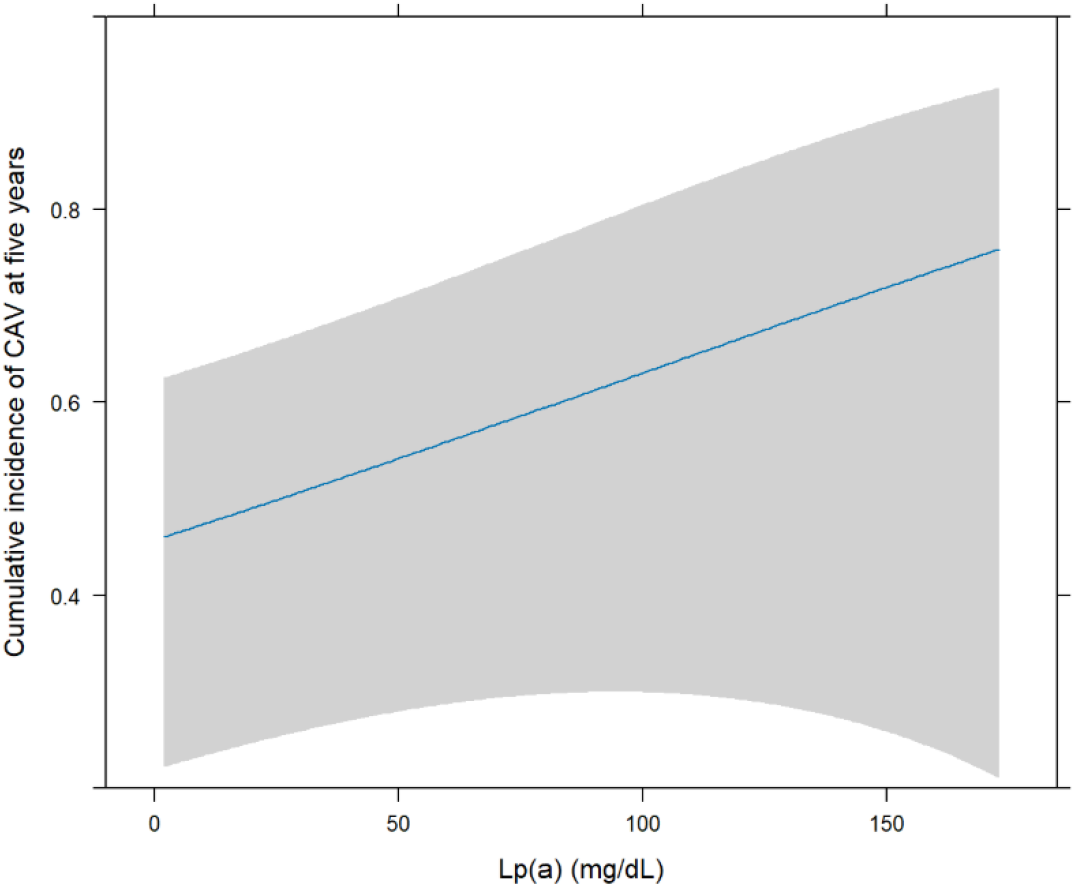
Model estimated CAV cumulative incidence at five years by Lp(a) after adjustment for confounders

As a sensitivity analysis, a Cox model with a categorical Lp(a) variable (i.e, Lp(a) ≥ 90 mg/dL vs Lp(a) < 90 mg/dL) was fitted after adjusting for the same covariates. The Cox model results found overall the relationship between Lp(a) and CAV was marginally significant (p=0.076). The HR comparing Lp(a) ≥ 90 mg/dL vs Lp(a) < 90 mg/dL was 1.51, 95% CI was 0.96 to 2.37, p=0.15. Among patients with LDL ≤ 100 mg/dL, after adjusting for the same covariates, Lp(a) was significantly related to the risk of developing CAV (HR=1.81, 95% CI was 1.09 to 2.99, p=0.022). The model-estimated CAV-free survival probability over time by Lp(a) categories is presented in **Figure 4**. However, the relationship between Lp(a) levels and risk of developing CAV was not significant among patients with LDL > 100 mg/dL. HR was 0.79, 95% CI was 0.21 to 2.99, p=0.733.

**Figure 4.**
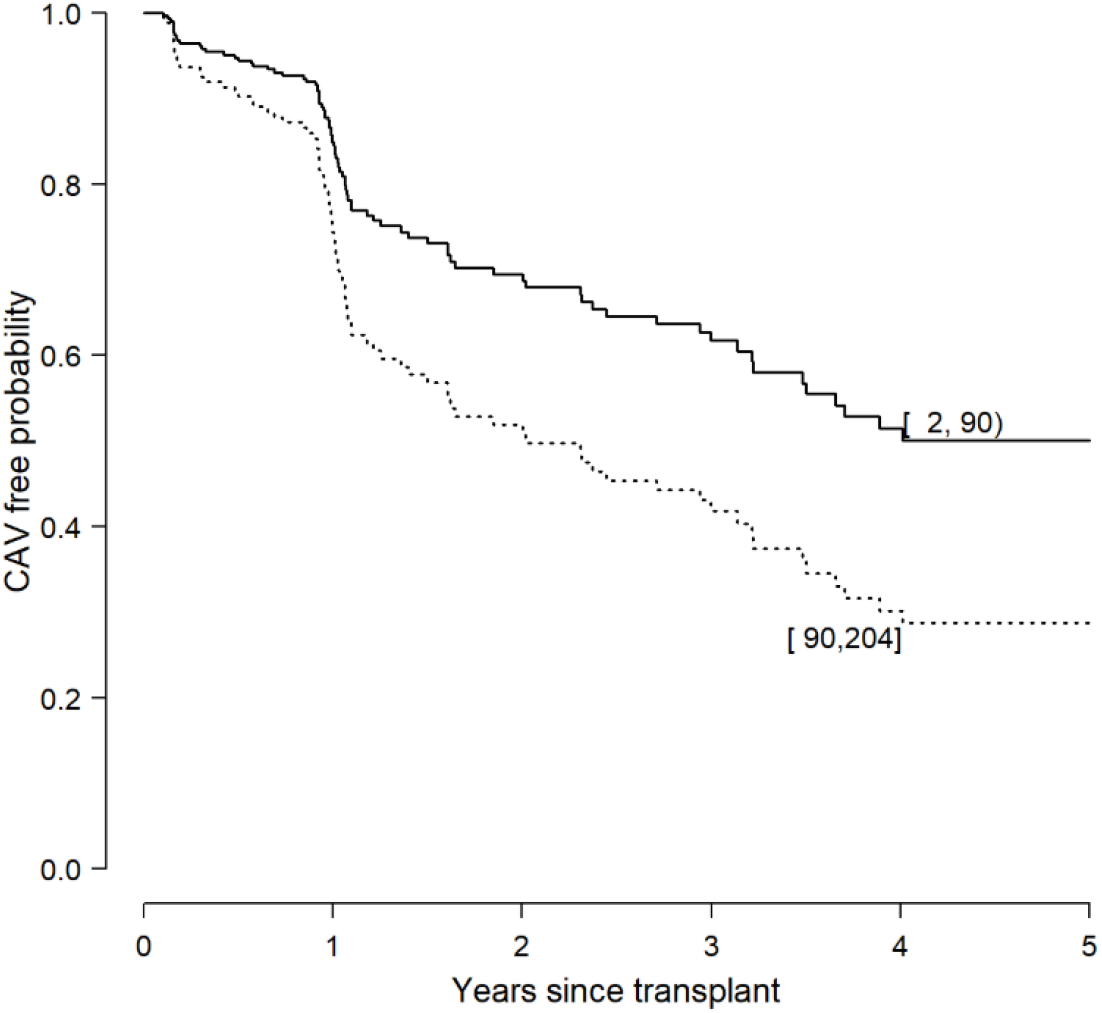
Model-estimated CAV-free survival probability over time stratified by Lp(a) ≥ 90 mg/dL or < 90 mg/dL

**Figure 5.**
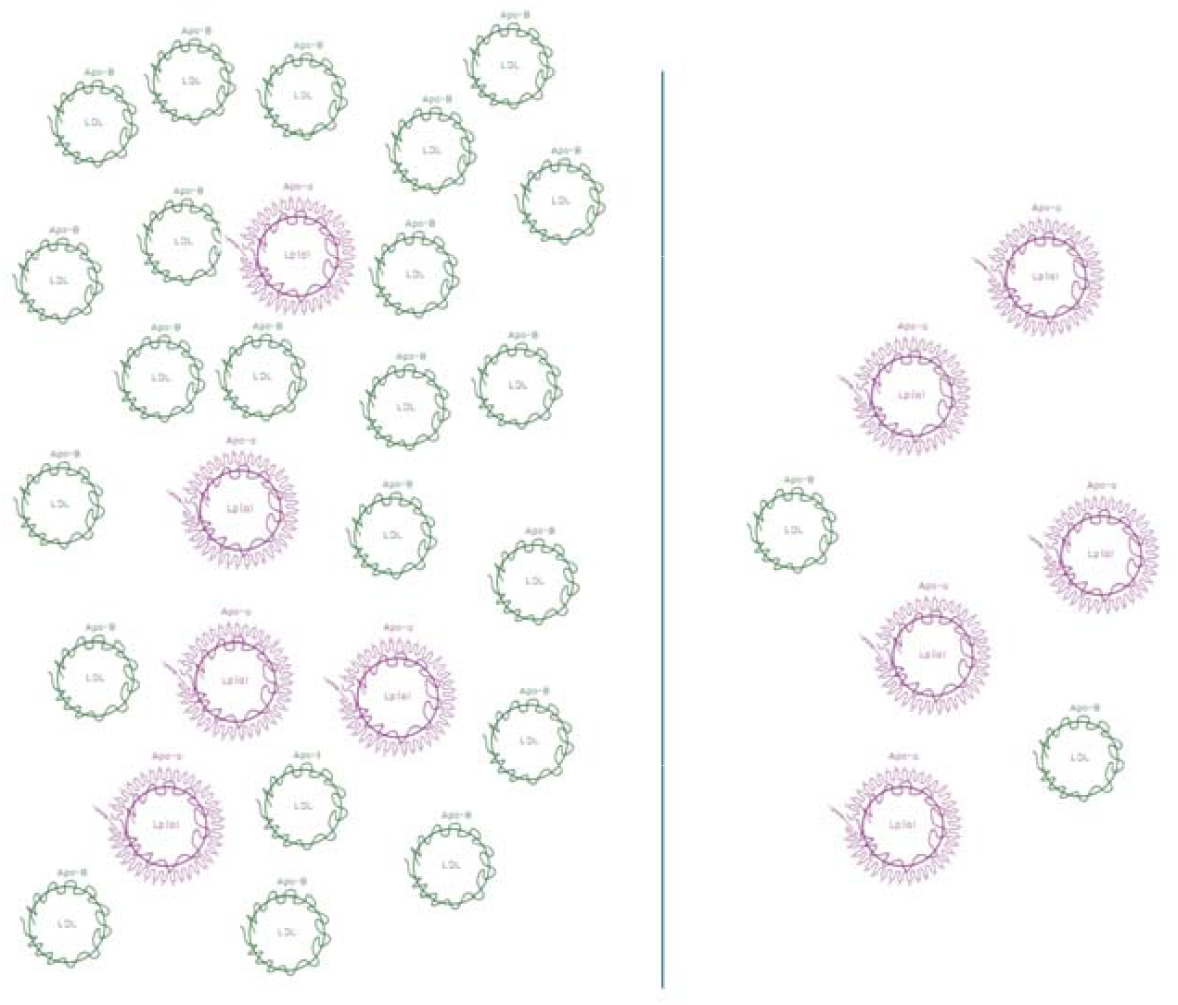
Schematic of possible outsize Lp(a) contribution to measured LDL levels at lower LDL values, on right, versus at higher LDL levels, on left

## Discussion

Among adult heart transplant recipients at our center, Lp(a) ≥ 90 mg/dL was associated with development of CAV, particularly at LDL ≤ 100 mg/dL. Because the cholesterol component of Lp(a) contributes to measured LDL, at lower LDL levels higher Lp(a) may have an outsize effect. This may be particularly important in transplant patients, for whom early initiation of statins and close management of dyslipidemia is standard of care^7^. There are few data for specific LDL targets in heart transplant recipients. However, the ISHLT 2023 guidelines suggest targeting LDL levels below 100 mg/dL, with more aggressive targets in patients with known CAV^7^. Indeed, among our patients, overall LDL levels were well-controlled, with mean and median LDL levels in both CAV-positive and negative patients in the 70 mg/dL range and lower The relatively low LDL levels in our cohort further supports the utility of using Lp(a) for CAV risk stratification in the transplant population. In addition, our work also suggests that lower Lp(a) levels < 90 mg/dL are associated with CAV-free survival in the lower LDL population.

Lp(a)’s structural differences may contribute to its role in CAV. Prior work has demonstrated that Lp(a) can stimulate growth of smooth muscle cells and selectively binds to the arterial wall via binding of the apo(a) subgroup to the extracellular matrix^14,16^. Lp(a) is also pro-inflammatory and has been shown to attract monocytes, increase cytokine expression, and affiliate with oxidized phospholipids that can increase the immune response^14,16^.

Importantly, standard lipid modifying therapies beyond lipoprotein apheresis do not lower Lp(a) sufficiently to reduce adverse cardiovascular events in the general population, and some work has found that statins may minimally increase Lp(a) levels^9^. Recent work has suggested that PCSK9 inhibitors lower Lp(a) and potentially may reduce cardiovascular events^21,22^. Some work has found that in patients with LDL around 70 mg/dL, PCSK9 inhibitors further lowered rates of cardiovascular events in patients with even mildly elevated Lp(a), further supporting the idea that Lp(a) may become a stronger contributor to residual cardiovascular risk at lower LDL levels^23^. There are multiple clinical trials currently examining other Lp(a) lowering drugs, including novel RNA based treatments. As additional Lp(a) lowering therapeutics become available, outcome trials in heart transplant patients will be essential to assess for efficacy in this subpopulation.

### Limitations

This study has several limitations. The majority of our patients had well-controlled LDL levels and the total number of patients with LDL > 100 mg/dL in both Lp(a) subgroups was relatively small, particularly as time progressed. In addition, this work was conducted at a single center. Patient demographics and practice patterns in terms of lipid and CAV monitoring and intervention may vary at other centers. Post-transplant monitoring was also not uniform for all patients, particularly during pandemic years. However, this was accounted for as best as possible through statistical analysis. We also defined CAV without including intravascular ultrasound grading, potentially reducing sensitivity of CAV detection. Finally, we faced the limitations inherent in chart review and certain variables not available in the chart were not included.

## Conclusion

At a high-volume heart transplant center, Lp(a) ≥ 90 mg/dL was associated with development of angiographic CAV among adult HT recipients, particularly at LDL ≤100 mg/dL. In patients with LDL ≤ 100 mg/dL, Lp(a) was also associated with CAV-free survival. Lp(a) may be a useful biomarker for risk stratification in transplant patients and a potential future target for CAV prevention with emerging Lp(a) therapeutics.

## Data Availability

All data produced in the present work are contained in the manuscript

## Supplemental Data

Among 154 patients with both pre- and post-transplant Lp(a) values, we averaged the pre- and post-transplant Lp(a) values and compared them to determine if there was any significant difference. Average post-transplant Lp(a) values were significantly higher than pre-transplant Lp(a) values (46.7 ± 48.2 mg/dL versus 21.0 ±43.4 mg/dL respectively, p=0.003). We also examined the overall trajectory of Lp(a) over time, and found the overall trajectory was not significantly changed (p=0.217).

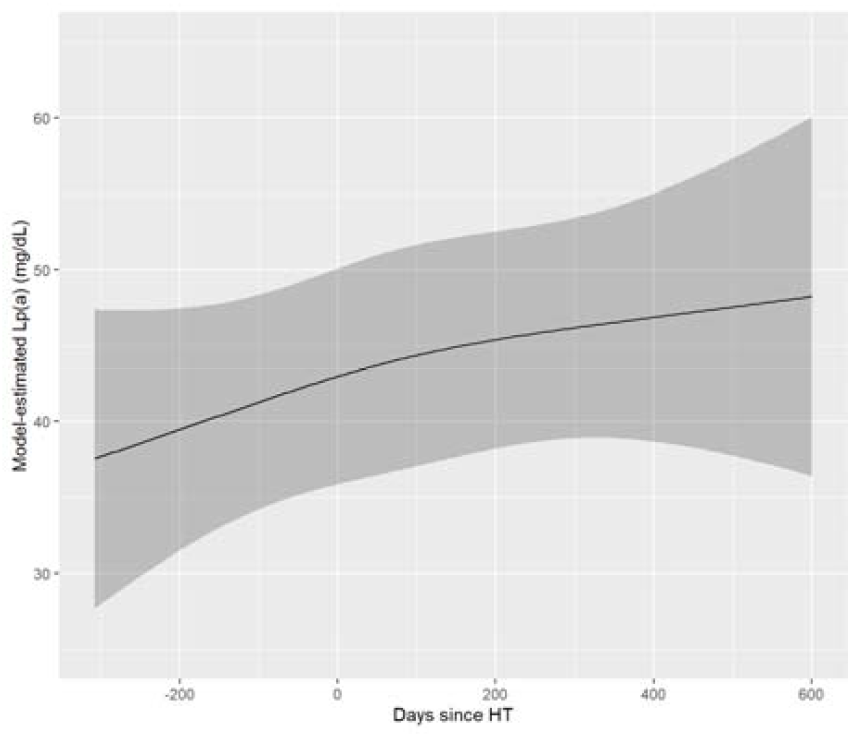

